# Surface sampling for SARS-CoV-2 in workplace outbreak settings in the UK, 2021-22

**DOI:** 10.1101/2023.02.17.23286079

**Authors:** Ian Nicholls, Antony Spencer, Yiqun Chen, Allan Bennett, Barry Atkinson

**Author notes:** Correspondence: Barry Atkinson.

## Abstract

**Aims:** To utilise environmental surface sampling to evaluate areas of SARS-CoV-2 contamination within workplaces to identify trends and improve local COVID-control measures.

**Methods and Results:** Surface sampling was undertaken at 12 workplaces that experienced a cluster of COVID-19 cases in the workforce between March 2021 and March 2022. 7.4% (61/829) of samples collected were positive for SARS-CoV-2 RNA by qPCR with only 1.8% (15/829) of samples identified with crossing threshold (Ct) values below 35.0. No sample returned whole genome sequence inferring RNA detected was degraded.

**Conclusions:** Few workplace surface samples were positive for SARS-CoV-2 RNA and positive samples typically contained low levels of nucleic acid. Although these data may infer a low probability of fomite transmission or other forms of transmission within the workplace, Ct values may have been lower at the time of contamination. Workplace environmental sampling identified lapses in COVID-control measures within individual sites and showed trends through the pandemic.

**Significance and Impact of the Study:** Prior to this study, few published reports investigated SARS-CoV-2 RNA contamination within workplaces experiencing cases of COVID-19. This report provides extensive data on environmental sampling identifying trends across workplaces and through the pandemic.

## INTRODUCTION

Severe acute respiratory syndrome coronavirus 2 (SARS-CoV-2) is the etiological cause of coronavirus disease 2019 (COVID-19) which emerged in Wuhan, China in late 2019. By March 2020, SARS-CoV-2 had spread globally and was declared a pandemic by the World Health Organisation (WHO, 2020).

The UK government’s initial COVID-19 control measures included limiting social interactions and substantially reducing occupancy within non-essential workplaces. While these measures limited potential exposure to SARS-CoV-2, the scope of workplaces deemed as essential meant millions of people were required to attend their place of work with an estimated 30% of adult workers required to travel to their place of work during the first UK lockdown period in May 2020 rising to 50% by May 2021 (ONS, 2021). These key-workers were found to have had experienced a greater exposure to, and be at increased risk of COVID-19 infection (Brown, Coventry & Pepper, 2021; Topriceanu *et al*., 2021).

While the majority of workplace COVID-19 outbreaks were associated with short- and long-term care facilities including hospitals, numerous outbreaks were also reported in non-care sectors including manufacturing, retail, education and hospitality (European Centre for Disease Prevention and Control, 2020; Chen *et al*., 2022; Hosseini *et al*., 2022). Congregate settings where workers are frequently in close proximity were notably affected with early reports from industries such as meat processing facilities and correctional institutes (Dyal *et al*., 2020; Saloner *et al*., 2020); however, outbreaks were reported in all workplace sectors.

A core component of understanding transmission risk within the workplace is identifying areas with SARS-CoV-2 contamination. These data not only identify higher risk areas or places where infection control measures require strengthening, but also highlight potential transmission risk areas through fomite transmission. Studies have shown that SARS-CoV-2 can remain viable on surfaces for numerous days after deposition (Riddell *et al*., 2020; van Doremalen *et al*., 2020; Paton *et al*., 2021; Sun *et al*., 2022) while the nucleic acid can be detected for numerous weeks (Liu *et al*., 2021a, 2021b; Paton *et al*., 2021; Coil *et al*., 2022). The persistence of SARS-CoV-2 viral particles and nucleic acid on surfaces therefore allows for sampling of workplaces to determine potential risk areas and transmission risk among workers.

As part of the COVID-19 Outbreak investigation to Understand Transmission study (COVID-OUT) study, environmental surface sampling was conducted at workplaces reporting cases of COVID-19 within their workforce to establish areas of contamination within workplaces, potential routes of transmission, and to learn lessons from workplace outbreaks which can be incorporated into wider guidance. The data generated could also provide insight into whether COVID-19 cases affecting businesses are predominately workplace-related (resulting from factors within the work environment) or workforce-related resulting from interactions outside of a place of work and beyond the control of the employer.

## METHODS

### Recruitment to the COVID-OUT study

Workplaces in the UK are required to report potential outbreaks to the Health and Safety Executive (HSE) and United Kingdom Health Security Agency (UKHSA). Identification and recruitment of workplaces experiencing cases of COVID-19 in their workforce was conducted using these databases as part of the wider COVID-OUT study (Chen *et al*., 2021). To be eligible for inclusion in this study, a workplace was required to have an attack rate of ≥ 5% at the time of notification in a workforce of ≥ 100 workers as well as qualifying as one of the following facilities: food processing plants, general manufacturing facilities, packaging and distribution centres, or large office buildings. Smaller workplaces (<100 workers) were approached if >5 workers were infected at the time of notification with projected increases. Workplaces meeting the criteria for study inclusion were approached for approval to participate in the study including the option for environmental surface sampling.

In addition, two control sites were sampled at separate times during the course of the study to represent the background levels of surface contamination which could be expected from a workplace which was not experiencing, and had not recently experienced, an elevated level of COVID-19 infections.

### Surface sampling

Environmental surface sampling was performed as soon as practicable after a participation agreement was in place. An on-site reconnaissance of the overall work environment was carried out to identify priority areas for sampling such as door handles, toilets, canteens, high-occupancy workstations/desks and locker rooms. Areas occupied by workers who had been recently diagnosed with COVID-19 infection were also targeted.

Surface sampling was carried out using either Blue Stick Sponge Swabs (Technical Service Consultants, TS/15-SH) or Blue Sponge Swabs (Technical Service Consultants, TS/15-B) for flat surfaces, and Universal Transport Medium (UTM) Swabs (Copan, 366C) for smaller or more detailed surface areas such as door handles. Where possible, samples were collected from an approximate 10cm x 10cm area to allow for estimation of contamination should heavily contaminated environments be identified. For each sample, an estimate of the area sampled was recorded in addition to the type of environment sampled and whether the sample was from a ‘high-touch’ area (likely to be contacted at least twice per working day by at least two separate individuals).

Sample team members wore an IRII surgical mask and two pairs of nitrile gloves to prevent potential (cross-)contamination of samples in addition to any other personal protective equipment required to operate in the specific work environment e.g., ear defenders. Collected samples were packaged into a UN3373 rated container for transport back to the laboratory and stored at 4°C until subsequent processing (16-60hrs).

### Sample Analysis

Samples were processed within a class II microbiological safety cabinet. Sponge samples were manually massaged by hand for approximately 30 seconds in their sample bags to homogenise the sample and release absorbed buffer which was then removed from the bag using a serological pipette and stored in a 2mL Sarstedt tube. UTM swabs were pressed against the inside edge of the collection tube in a rolling motion to release any retained UTM which was then removed by pipette and stored in a 2mL Sarstedt tube. When lysis and RNA purification was not carried out immediately, samples were placed into storage in a -80°C freezer.

SARS-CoV-2 RNA was extracted and purified from samples using QIAamp Viral RNA Mini Kit (Qiagen, 52906) following the manufacturer’s centrifuge protocol. Quantitative PCR (qPCR) was carried out using the Viasure SARS-CoV-2 detection kit (CerTest, VS-NC02) following manufacturer’s instructions. The targets for this assay were the SARS-CoV-2 nucleocapsid gene (N) and polyprotein open reading frame 1a and 1b (ORF1ab). Assay analysis was performed on a QuantStudio 5 thermocycler (Applied Biosystems) against an in-house N gene standard curve to determine approximate genome copy number in positive samples. The cycling conditions were as follows: one reverse transcription cycle of 45°C for 15 mins followed by denaturation at 95°C for 2 mins, then 45 cycles of 95°C for 10 secs and 60°C for 50 secs with quantification of fluorescence performed at the end of 60°C step. All samples were tested in duplicate against both targets. Samples were deemed positive when both duplicate tests returned a valid crossing threshold (Ct) value for either target. If only a single replicate returned a valid Ct value, repeat analysis was performed; if the same result was returned the sample was referred to as ‘suspected positive’ unless two valid Ct values were recorded for the other target. Samples were deemed negative when no Ct value was returned for either replicate in both targets. Invalid Ct values were those above Ct 38.0 which is the upper limit described by the kit manufacturer; samples with Ct values above 38.0 were deemed as ‘technical negatives’ and considered as negatives.

### Additional analysis

Samples returning Ct values of ≤ 35.0 cycles were subjected whole genome sequencing using the ARTIC Network protocol (Quick, 2020, using v4.1 primers) to elucidate detailed genetic composition of the contaminating virus. Any sample returning complete viral genome would be eligible for viral isolation to demonstrate the presence of infectious virus.

## RESULTS

### Workplace recruitment

Environmental surface sampling for SARS-CoV-2 contamination was performed at 12 workplaces (Sites 001-005, 007-010, 012-013 and 020) and two control sites (C1 and C2) between March 2021 and March 2022 (Table 1). The recruited sites spanned numerous essential workplace sectors including manufacturing (four sites), food (four sites), office-based workplaces (two sites), distribution (one site) and critical infrastructure (one site); the two control sites were both within the critical infrastructure sector. Cases of COVID-19 among the workforce of recruited sites had either ceased or were declining in all cases at the time of environmental sampling; however, cases of COVID-19 were still being reported among the workforce at sites 002, 003, 005 and 008 when sampling was performed. Sites 001, 002, 003, 004, 005, 010, 012 and 013 met the inclusion criteria of ≥ 100 employees and ≥ 5% attack rate at the time of the outbreak report; sites 008, 009 and 020 had ≤ 100 staff but ≥ 5 confirmed cases at the time of the outbreak report with projected increases. Site 005 did not achieve a ≥ 5% attack rate in its workforce of more than 1000 employees; however, this site recorded a large number of cases in a short period of time and was recruited based on projected increases. The two control sites both had ≥ 300 staff but without indication of a potential outbreak within the workplace; however, both sites had sporadic cases at the time of sampling.

**Table 1:** Details of workplaces sampled for the presence of SARS-CoV-2 RNA as part of the COVID-OUT study. Sites 006, 011 and 014-019 did not give approval (or were not eligible) for the environmental sampling component of the study. C1 = control site 1, C2 = control site 2, NA = not applicable, Unknown = no outbreak reported but sporadic cases likely present in workforce. †Workforce size is presented as a range as this is potentially identifiable information. ‡Attack rate calculated as the number of COVID cases reported in the outbreak report divided by exact workforce size at that specific site (this is does distinguish between staff working on site and those who may be working from home during the outbreak period). *Outbreak confined to one cohort within the work environment; data provided for both the cohort and the wider workplace. **Last case reported at the time of sampling; however, site was closed for nine days prior to sampling with minimal staff present.

| Site | Workplace Sector | Workforce size<sup>†</sup> | Estimated attack rate<sup>‡</sup> | Sampling Performed | Time from outbreak cases to sampling |
| --- | --- | --- | --- | --- | --- |
| 001 | Manufacturing | 200-300 | 10% | March 2021 | >10 days |
| 002 | Manufacturing | 200-300 | 14% | March 2021 | Ongoing |
| 003 | Distribution | >1000 | 6% | May 2021 | Ongoing |
| 004 | Office | 100-200 | 12% | May 2021 | >14 days |
| 005 | Food | >1000 | 3% | June 2021 | Ongoing |
| 007 | Critical Infrastructure | 100-200 (20-30)* | 5% (39%) | July 2021 | >14 days |
| 008 | Office | <50 | 55% | September 2021 | Ongoing** |
| 009 | Manufacturing | <50 | 14% | October 2021 | >21 days |
| 010 | Manufacturing | 200-300 | 10% | November 2021 | >7 days |
| 012 | Food | 200-300 | 9% | January 2022 | >14 days |
| 013 | Food | 100-200 | 21% | February 2022 | >21 days |
| 020 | Food | <50 | 32% | March 2022 | 4 days |
| C1 | Critical Infrastructure | 500-1000 | Unknown | July 2021 | NA |
| C2 | Critical Infrastructure | 300-400 | Unknown | March 2022 | NA |

The complex recruitment process resulted in several sites being sampled at least two weeks from the last reported case in the work environment linked to the reported outbreak (sites 004, 007, 009, 012 and site 013).

### Detection of SARS-CoV-2 RNA across workplaces

A total of 829 samples were collected from the 12 recruited workplaces (average = 63.7 samples per site). 723 samples (87.2%) were negative for SARS-CoV-2 RNA; 61 samples (7.4%) were identified as positive for SARS-CoV-2 RNA and 45 samples (5.4%) identified as suspected positive (Table 2). Only 15/829 (1.8%) of samples collected across the 12 workplaces returned Ct values of ≤ 35.0. Estimated genome copy numbers for samples with Ct values of ≤ 35.0 ranged from 6.8×10^2^ to 6.6×10^3^ genomic copies/cm ^2^.

**Table 2:** Sampling results from recruited workplaces reporting a recent outbreak of COVID-19 in their workforce. Type of site indicated using the following codes: (M) Manufacturing, (D) Distribution, (O) Offices, (F) Food sector industry, or (CI) Critical infrastructure. Ct = Crossing threshold value based on average of two duplicate samples against the nucleocapsid gene.

| Site (Month) | Samples collected | Positive (%) | Suspected (%) | Negative (%) | Ct ≤35.0 (%) |
| --- | --- | --- | --- | --- | --- |
| 001 (M) | 36 | 14 | 3 | 19 | 2 |
| March 2021 |  | (38.9%) | (8.3%) | (52.8%) | (5.6%) |
| 002 (M) | 66 | 8 | 11 | 47 | 0 |
| March 2021 |  | (12.1%) | (16.7%) | (71.2%) | (0.0%) |
| 003 (D) | 76 | 25 | 14 | 37 | 7 |
| May 2021 |  | (32.9%) | (18.4%) | (48.7%) | (9.2%) |
| 004 (O) | 69 | 2 | 7 | 60 | 0 |
| May 2021 |  | (2.9%) | (10.1%) | (87.0%) | (0.0%) |
| 005 (F) | 60 | 1 | 6 | 53 | 1 |
| June 2021 |  | (1.7%) | (10.0%) | (88.3%) | (1.7%) |
| 007 (CI) | 90 | 0 | 0 | 90 | 0 |
| July 2021 |  | (0.0%) | (0.0%) | (100.0%) | (0.0%) |
| 008 1<sup>st</sup> visit (O) | 60 | 10 | 1 | 49 | 5 |
| September 2021 |  | (16.7%) | (1.7%) | (81.7%) | (8.3%) |
| 008 2<sup>nd</sup> visit (O) | 42 | 1 | 1 | 40 | 0 |
| September 2021* |  | (2.4%) | (2.4%) | (95.2%) | (0.0%) |
| 009 (M) | 70 | 0 | 1 | 69 | 0 |
| October 2021 |  | (0.0%) | (1.4%) | (98.6%) | (0.0%) |
| 010 (M) | 55 | 0 | 0 | 55 | 0 |
| November 2021 |  | (0.0%) | (0.0%) | (100.0%) | (0.0%) |
| 012 (F) | 65 | 0 | 0 | 65 | 0 |
| January 2022 |  | (0.0%) | (0.0%) | (100.0%) | (0.0%) |
| 013 (F) | 69 | 0 | 0 | 69 | 0 |
| February 2022 |  | (0.0%) | (0.0%) | (100.0%) | (0.0%) |
| 020 (F) | 71 | 0 | 1 | 70 | 0 |
| March 2022 |  | (0.0%) | (1.4%) | (98.6%) | (0.0%) |
| Total | 829 | 61 | 45 | 723 | 15 |
|  |  | (7.4%) | (5.4%) | (87.2%) | (1.8%) |

A total of 136 samples were collected from two control sites (average = 68 per site) that were not currently experiencing an outbreak of SARS-CoV-2. 131 samples (96.4%) were negative for SARS-CoV-2 RNA and one sample (0.7%) was identified as positive with another four samples (2.9%) identified as suspected positive (Table 3). The lowest Ct value identified at the control sites was 36.9; estimated genome copy numbers for the five samples returning a Ct value ranged from 1.1×10^2^ to 1.2×10^3^ genomic copies/cm ^2^.

**Table 3:**
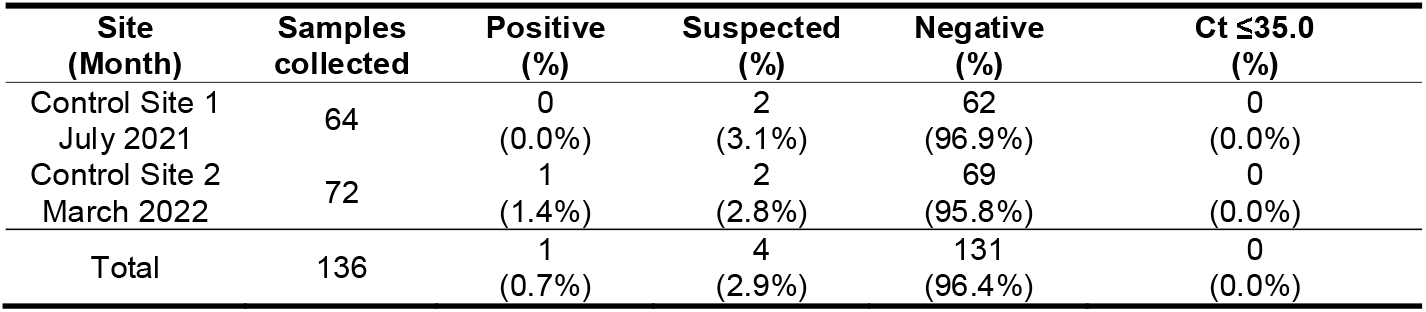
Sampling results from two critical infrastructure control sites; neither site reported a recent outbreak of COVID-19 within their workforce. Ct = Crossing threshold value based on average of two duplicate samples against the nucleocapsid gene.

| Site (Month) | Samples collected | Positive (%) | Suspected (%) | Negative (%) | Ct ≤35.0 (%) |
| --- | --- | --- | --- | --- | --- |
| Control Site 1 July 2021 | 64 | 0 (0.0%) | 2 (3.1%) | 62 (96.9%) | 0 (0.0%) |
| Control Site 2 March 2022 | 72 | 1 (1.4%) | 2 (2.8%) | 69 (95.8%) | 0 (0.0%) |
| Total | 136 | 1 (0.7%) | 4 (2.9%) | 131 (96.4%) | 0 (0.0%) |

As shown in Figure 1, samples collected at the first three workplaces (sites 001-003; March – early May 2021) had a high proportion of positive samples (12.1-38.9%). In contrast, from late May 2021 onwards (Sites 004-020), only one site was identified with >3% of positive samples. A similar trend was seen with samples identified as ‘suspected positive’ with 8.3-18.4% of samples collected at the first five sites (March-June 2021) whereas no site was identified with more than 2.4% of samples as suspected positive after June 2021. The outlier in terms of positive samples was the first of two visits to Site 008 (September 2021) where 16.7% of samples were positive; this investigation was conducted at the start of a rise in community cases associated with the Delta variant in a workplace with a 55% attack rate among the workforce (Atkinson *et al*., 2022).

**Figure 1:**
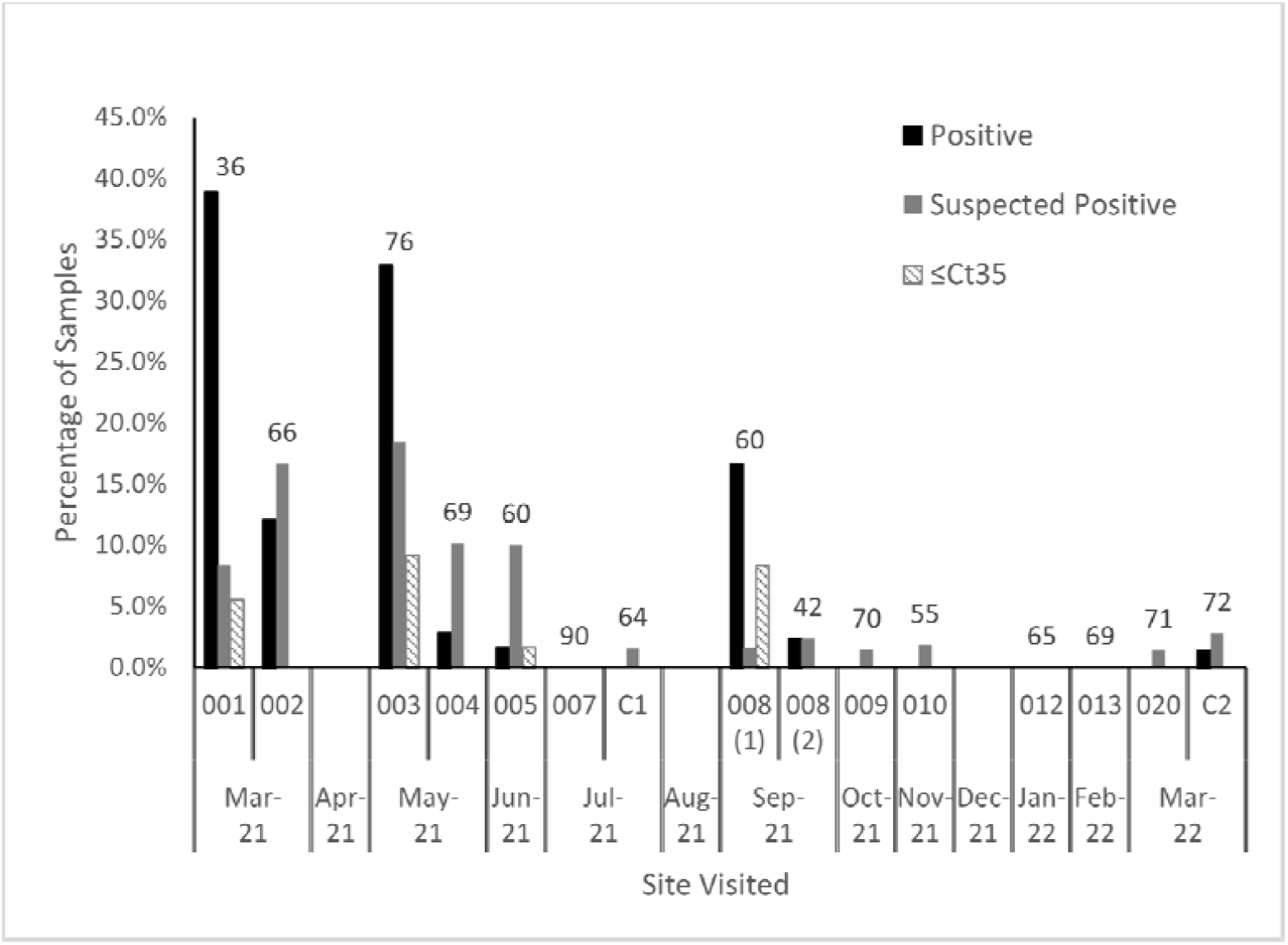
Proportion of positive samples, suspected positive samples and samples returning Ct values of ≤ 35.0 by site visited.

Only 15/829 (1.8%) of samples collected at recruited workplaces returned Ct values of ≤ 35.0. These 15 samples were identified at four distinct sites: 001 (n=2), 003 (n=7), 005 (n=1) and 008 (n=5).

No positive or suspected positive samples were found at four of the 12 workplaces sampled (007, 010, 012 and 013), and in the final six months of the study (sites 009-010, 012-013 and 020) no positive samples (0/330; 0.0%) and only two suspected positive samples (2/330; 0.6%) were identified.

### Detection of SARS-CoV-2 RNA within workplaces

SARS-CoV-2 RNA was detected in nearly all types of workplace environment (Figure 2). The highest rate of confirmed positivity was observed in locker rooms (13.4%) followed by general work areas (8.8%), ventilation (6.9%), canteens (4.2%) and toilets (2.0%). No positive samples were identified in corridors or from samples collected outside.

**Figure 2:**
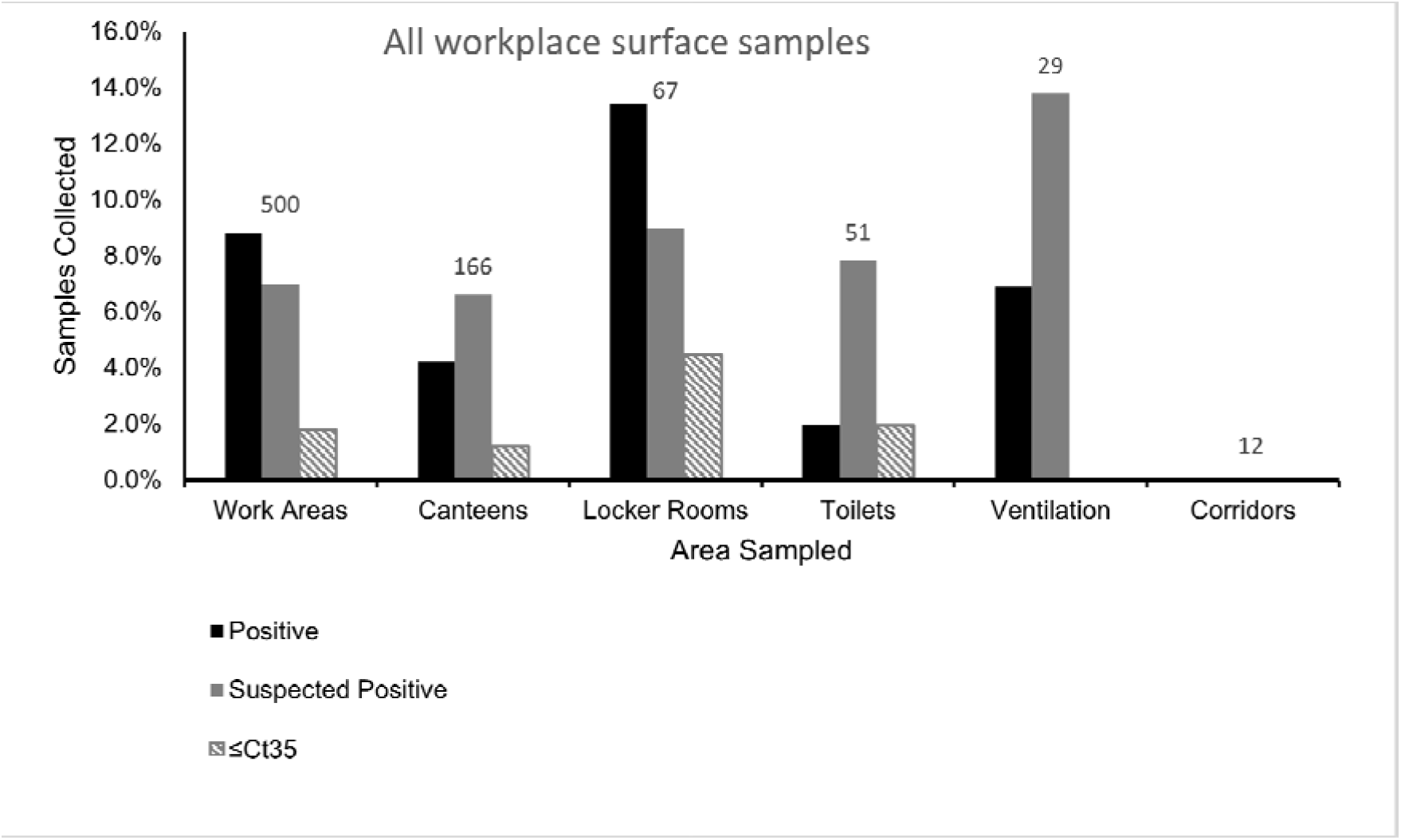
Proportion of positive samples, suspected positive samples and samples returning Ct values of ≤ 35.0 from 829 surface samples collected from recruited workplace ordered by area of sampling. Four samples, which included one suspected positive, collected from outdoor locations are not shown for y-axis scaling purposes.

The level of suspected positive samples was broadly similar for work areas, canteens, locker rooms and toilets (6.6-9.0%). The level of suspected positives was higher for ventilation (13.8%) and for outside samples (25.0%); however, only four samples were collected from outside environments. No suspected positive samples were identified from corridors.

Samples with qPCR Ct values ≤ 35.0 cycles were only identified in locker rooms (4.5%), toilets (2.0%), work areas (1.8%) and canteens (1.2%).

524 of the 829 samples collected from recruited workplaces were classified as being from ‘high-touch’ locations (Figure 3). Levels of confirmed positivity were broadly similar for high-touch samples versus all samples from that environment for work areas (7.5% vs 8.8%), canteens (4.5% vs 4.2%) and toilets (2.9% vs 2.0%); however, there was a small reduction in the positivity rate for high-touch samples in locker rooms (9.1% vs 13.4%) and no positive samples were identified from high-touch ventilation samples (0.0% vs 6.9%). As there were no positive samples identified in any corridor or outside sample, none of the high-touch samples in these groups were positive.

**Figure 3:**
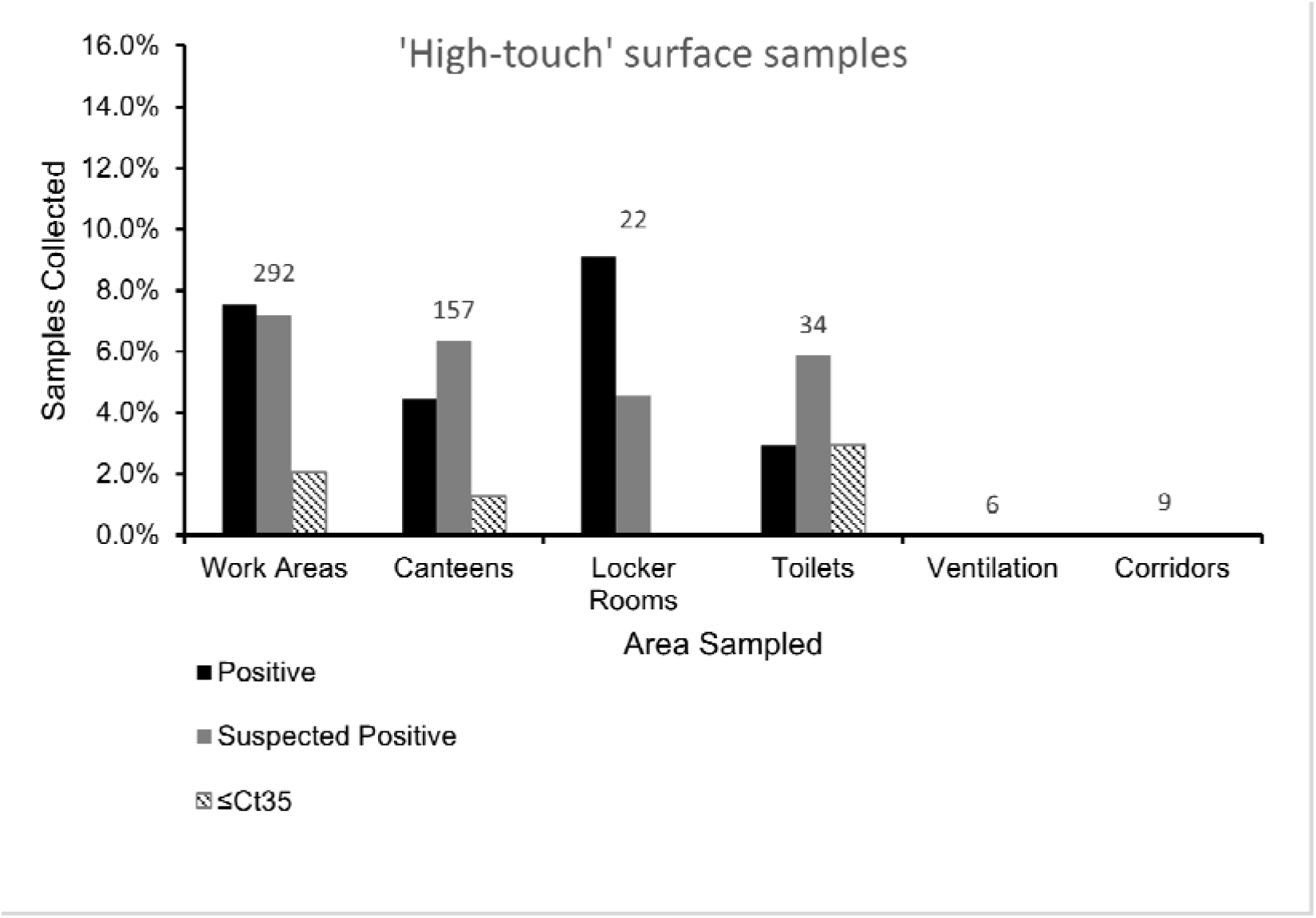
Proportion of positive samples, suspected positive samples and samples returning Ct values of ≤ 35.0 from 524 surface samples classed as ‘high-touch’ locations collected from recruited workplace ordered by area of sampling. Four samples, which included one suspected positive, collected from outdoor locations are not shown for y-axis scaling purposes.

A similar trend was seen in suspected positive samples with comparable levels for high-touch samples versus all samples in that environment for outside samples (25.0% vs 25.0%), work areas (7.2% vs 7.0%), and canteens (6.4% vs 6.6%). There was an apparent reduction in suspected positivity rate in high-touch samples for toilets (5.9% vs 7.8%), locker rooms (4.5% vs 9.0%) and ventilation samples (0.0% vs 13.8%). As there were no suspected positive samples identified in any corridor sample, none of the high-touch samples in this group was suspected positive.

For samples with a Ct value ≤ 35.0, levels were marginally higher for high-touch samples versus all samples in that environment for toilets areas (2.9% vs 2.0%), work areas (2.1% vs 1.8%) and canteens (1.3% vs 1.2%). No Ct values ≤ 35.0 were identified in high-touch locker room samples compared to 4.5% of all samples with Ct values ≤ 35.0 within this environment. As there were no samples with Ct values ≤ 35.0 identified in any ventilation, corridor, or outside sample, none of the high-touch samples in these groups had Ct values ≤ 35.0.

### Whole genome sequence analysis

The 15 samples returning N gene Ct values of ≤ 35.0 (32.7 to 35.0) were subjected to whole genome sequence analysis. No sample yielded complete genome data and only two samples generated any contiguous sequence once quality control standards were applied. Approximately 55% of the SARS-CoV-2 genome was recovered from a sample collected from a window handle at Site 008; however, the longest contiguous sequence was ~1300 bases implying the nucleic acid was sheared potentially through degradation in the environment from the contamination event until the point of sampling (>9 days). The other sample to return sequence data was collected in a toilet at Site 005; however, <500 bases were called representing just over 2% of the viral genome.

## DISCUSSION

Workplace outbreaks of COVID-19 were common across multiple workplace sectors during the pandemic despite significant efforts including government guidance and frameworks aimed at reducing the spread of infection. While government guidance and workplace COVID-control measures were aimed at reducing transmission of COVID-19 within the workplace, the effect of non-workplace transmission among the workforce is difficult to elucidate. Workforce mixing can occur in non-workplace locations such as social locations, transport to and from the workplace, and within the home. This study aimed to investigate areas of surface contamination within workplaces which experienced a recent COVID-19 outbreak and use this data to inform improvements to control measures which can mitigate reoccurrence.

Between March 2021 and March 2022, 829 surface samples were collected from 12 workplaces that recently experienced an outbreak of COVID-19 among their workforces. These workplaces were located across the United Kingdom spanning multiple business sectors and encompassing a variety of workforce sizes. Across all sites visited, only 7.4% of samples collected were identified as positive by qPCR analysis with a further 5.4% identified as ‘suspected positive’ likely indicating SARS-CoV-2 genetic material close to the limit of detection for the assay. The remaining 87.2% of samples collected were negative. Estimates of genome copies per cm^2^ of SARS-CoV-2 RNA positive surface samples were calculated using a standard curve with no sample collected exceeding 6.6×10^3^ genomic copies/cm^2^. While finding limited amounts of SARS-CoV-2 nucleic acid at sites reporting a COVID-19 cases may seem counter-intuitive, low levels of contamination were reported in similar studies including the in areas recently occupied by symptomatic workers (Gholipour *et al*., 2020; Marshall *et al*., 2020; Mouchtouri *et al*., 2020; Cherrie *et al*., 2021; Marcenac *et al*., 2021; de Rooij *et al*., 2023) and in some clinical settings occupied by infected patients (Ryu *et al*., 2020; Goel *et al*., 2022; Warren *et al*., 2022). Two control sites were also sampled as part of this study; both positive and suspected positive samples were identified at these sites albeit at lower levels than in recruited sites. These findings are likely due to both control sites reporting COVID-19 cases near the time of sampling; however, the levels were below the inclusion criteria for the study.

Samples with Ct values less than 35.0 were detected only at four sites (sites 001, 003, 005, and 008) and in no samples collected after September 2021. The five samples with Ct values of ≤ 35.0 at Site 008 were identified after the apparent decrease in environmental surface sampling positivity rates seen in late May 2021 implying that a decrease in the proportion of samples identified as SARS-CoV-2 positive does not necessarily correlate with level of contamination that can be observed when positive samples are identified. The reduction in sample positivity levels after May 2021 and the absence of positive samples after September 2021 is likely due to a combination of factors. It is possible that one significant factor was the impact from the COVID-19 vaccine rollout starting in December 2020 with more than 34 million priming vaccine doses administered by May 2021 (GOV.UK, 2022). As vaccines were made available using a tiering system based on age and at-risk status, most working age individuals would likely have received at least one dose by summer 2021 when environmental sampling positivity rates declined. Some studies suggest that vaccine status has no effect on viral titre and shedding by infected individuals (Boucau *et al*., 2022; Riemersma *et al*., 2022); however, other studies suggest that infectious viral titre and/or shedding duration is reduced in vaccinated individuals (Ke *et al*., 2022; Plante *et al*., 2022; Puhach *et al*., 2022; Tian *et al*., 2022). Additionally, a ferret model study found a significant reduction in viral titre in both nasal wash and oral swabs amongst animals vaccinated with only a primer dose of Astra-Zeneca vaccine 7 days after viral challenge (Marsh *et al*., 2021). A reduction in viral titre and shortened shedding therefore may reduce environmental contamination from vaccinated cases.

Samples producing Ct values of ≤ 35.0 were submitted for whole genome sequencing; however, only two yielded sequence data and neither returned more than 55% of the genome. The inability to recover full genome sequence infers degradation of genomic material between the original deposition and environmental sampling. While this suggests the lack of infection-competent material in the samples collected, these contamination events may have harboured infectious material prior to sampling and does not conclusively rule out the potential for fomite transmission in the workplace.

Comparison of the types of surfaces sampled showed positives and suspected positives from all major surface types. Samples from locker rooms were the most common area to return a qPCR positive (13.8% of locker room samples) and a Ct value ≤ 35.0 (4.5% of locker room samples) indicative of higher amount of nucleic acid; this may reflect an increased risk due to being a small congregative area but could also reflect an oversight of local COVID control measures and cleaning regimens as most locker rooms did not have the same cleaning standards seen elsewhere in the workplaces. Toilets, canteens and general work areas had a comparably lower rate of positivity, and high-touch surfaces had comparable, or lower, rates of positivity in all environments; it is likely that enhanced cleaning regimens in high-use work areas contribute to these findings.

Data generated as part of this study provided site-specific information that was used to improve local COVID-19 control measures. A report of surface contamination and interpretation was provided to each site within five working days of sampling allowing for alterations to cleaning and infection control regimes locally if required. For example, sampling of Site 008 identified a Ct 33.6 sample from a window handle; after consultation, it was confirmed that window handles had been omitted from their enhanced cleaning procedures. Additionally, surface sampling showed an area with a high positivity rate; this area was operated by a contractor who had not updated the infection control policies in line with new guidance. These findings highlight the benefit that rapid environmental sampling can offer to workplaces experiencing outbreaks.

Although the data presented provide substantial information regarding workplace contamination, there are several limitations that require consideration. The time from reporting a potential workplace outbreak until environmental sampling could be performed was frequently more than 14 days due to site approval requirements of the study meaning some nucleic acid degradation may have occurred which would affect levels detected by qPCR. This delay also meant that air sampling of workplaces was not considered as part of the study. Additionally, while COVID-OUT was a research study, the lead organisations (UKHSA and HSE) have regulatory roles which may not only have resulted in a lower rate of participation in the study but could also have resulted in participating sites modifying the environment before the sampling team arrived as they may view the sampling visit as an official audit. Even if all sites sampled were genuine representations of the work environments at the time of cases being reported, the breadth of business and worker functions combined with epidemiological differences in local infection rates, the circulating SARS-CoV-2 variant, and increasing vaccination rates through the study period limits the conclusions that can be drawn when comparing individual sites to one another or on trends. Finally, assessment of contamination was made using qPCR Ct values; while this provide a semi-quantitative assessment, it does not distinguish between viable and degraded viral material, and variation can occur between reactions which has greater impact on accuracy near the limit of detection which was frequently the level of contamination observed in this study.

The environmental sampling carried out in this study provides data on the occurrence and the level of SARS-CoV-2 contamination seen in a variety of workplaces experiencing COVID-19 cases within their workforce throughout the pandemic. From a site-specific perspective, the data generated provided insight for where COVID control measures, workforce practices and regular cleaning may have been insufficient or may have lapsed; collectively, these results highlight common themes that may contribute to overall transmission either directly or as an indication of where transmission risks are the highest.

## Data Availability

All data produced in the present work are contained in the manuscript

## ACKOWLEDGMENTS

The authors wish to acknowledge the contributions of the UKHSA Genomics team for sequencing samples and from the COVID-OUT workplace recruitment team (Chris Keen, Gillian Frost, Joan Cook, Gary Dobbin, Derek Morgan, Vince Sandys, Matthew Coldwell, Andrew Simpson, Adam Clarke, Alice Graham, Hannah Higgins, **Christina Atchison and Helen Collins) for assistance with recruitment and completing the wider study**.

## CONFLICTS OF INTEREST

No conflict of interest declared.

## DISCLOSURE STATEMENT

The contents of this paper, including any opinions and/or conclusions expressed, are those of the authors alone and do not necessarily reflect Health and Safety Executive or UK Health Security Agency policy.

## FUNDING STATEMENT

This work was supported by funding from the PROTECT COVID-19 National Core Study on Transmission and Environment, managed by the Health and Safety Executive on behalf of HM Government.

## ETHICAL APPROVAL

The COVID-OUT study has been approved by the NHS North East Research Ethics Committee (Reference 20/NE/0282).

## Notes

### Competing Interest Statement

The authors have declared no competing interest.

